# Survey of symptoms following COVID-19 vaccination in India

**DOI:** 10.1101/2021.02.08.21251366

**Authors:** Rajeev Jayadevan, Ramesh Shenoy, Anithadevi TS

## Abstract

**Background:** COVID-19 vaccines have been rolled out recently in several parts of the world. Although the protective efficacy is frequently discussed, little is known about the real-world post-vaccination experience outside of clinical trial conditions. Knowledge about what to expect after vaccination will help educate the public, dispel misinformation and reduce vaccine hesitancy.

**Aim:**

1. To assess the immediate response to the first dose of COVID-19 vaccine.
2. To study the spectrum of post-vaccination symptom profile for individual vaccines.

**Methods:** A cross-sectional online survey was done which included questions pertaining to the immediate post vaccination experience in India.

**Results:** A total of 5396 people responded to the survey over a one-week period from 29 January to 4 February. Overall, 65.9 % of respondents reported at least one post-vaccination symptom. Tiredness (45%), myalgia (44%), fever (34%), headache (28%), local pain at injection site (27%), joint pain (12%), nausea (8%) and diarrhea (3%) were the most prevalent symptoms. The chance of having symptoms decreased with advancing age. The frequency of symptoms was 81% (3rd decade or 20-29 years), 80% (4th decade or 30-39 years), 68% (5th decade), 58% (6th decade), 45% (7th decade), 34% (8th decade) and 7% (9th decade, 80-90 years). Post-vaccination symptoms were more likely to be reported by women (74.7%) compared to men (58.6%) (p < 0.001). Among those who reported symptoms, 79% noticed them within the first 12 hours. 472 out of 5396 (8.7%) reported past history of COVID-19. Their symptom profile was not different to those who did not have a past history.

**Conclusions:** Two-thirds of healthcare professionals who completed the survey reported mild and short-lived post-vaccination symptoms. Tiredness, myalgia and fever were most commonly reported. These symptoms were consistent with an immune response commonly associated with vaccines, and correlated with the findings from previously published phase 2/3 trials. In 90% cases, the symptoms were either milder than expected or meeting the expectation of the vaccine recipient. No serious events were reported. Symptoms were more common among younger individuals. There was no difference in symptoms among those who had a past history of COVID-19.

## Background

COVID-19 vaccines have been rolled out recently in several parts of the world. Although the protective efficacy is frequently discussed, little is known about the real-world post-vaccination experience outside of clinical trial conditions. Knowledge about what to expect after vaccination will help educate the public, dispel misinformation and reduce vaccine hesitancy.

## Methods

A cross-sectional online survey was done which included questions pertaining to the immediate post vaccination experience in India. During the initial phase of vaccination in India launched on January 16, healthcare workers received the first dose as part of the priority list. The survey was sent for the attention of those who received COVID-19 vaccine till 3 February 2021.

Questions were formatted in binary fashion to the extent possible, with descriptive features added to the section on symptom profile. Provision was provided to add other outcomes or descriptions that further qualified the experience. The data presented in the study were exclusively obtained through the online survey.

### Statistical Analysis

Descriptive Statistics were used to assess the baseline characteristics of the data. All quantitative variables are presented as mean and standard deviation, and all qualitative variables in frequency and percentages. For the comparison of categorical variables, either chi square or fishers exact test were used. For continuous variables one-way ANOVA test/ Kruskal Wallis test were used. All the data were entered in Microsoft excel and analyzed using SPSS version 20.00.

## Results

A total of 5396 people responded to the survey over a one-week period from 29 January to 4 February.

Among those who responded to the survey, the majority were doctors (85.8%), followed by nurses (6.2%), technicians (1.1%) and others. This does not necessarily reflect the relative proportion of those who received the vaccine. 56% of the respondents were male, 44% were female.

Among the respondents, 5128 (95%) had received Covishield (Astra-Oxford vaccine manufactured by Serum Institute, India), 180 (3.3%) received Covaxin (Bharat Biotech, India), while 44 (0.8%) each had received Pfizer-Biontech and Sinopharm vaccine from other nations. Majority (98.3%) of the respondents had received either Covaxin or Covishield. (Figure 1)

**Figure 1:**
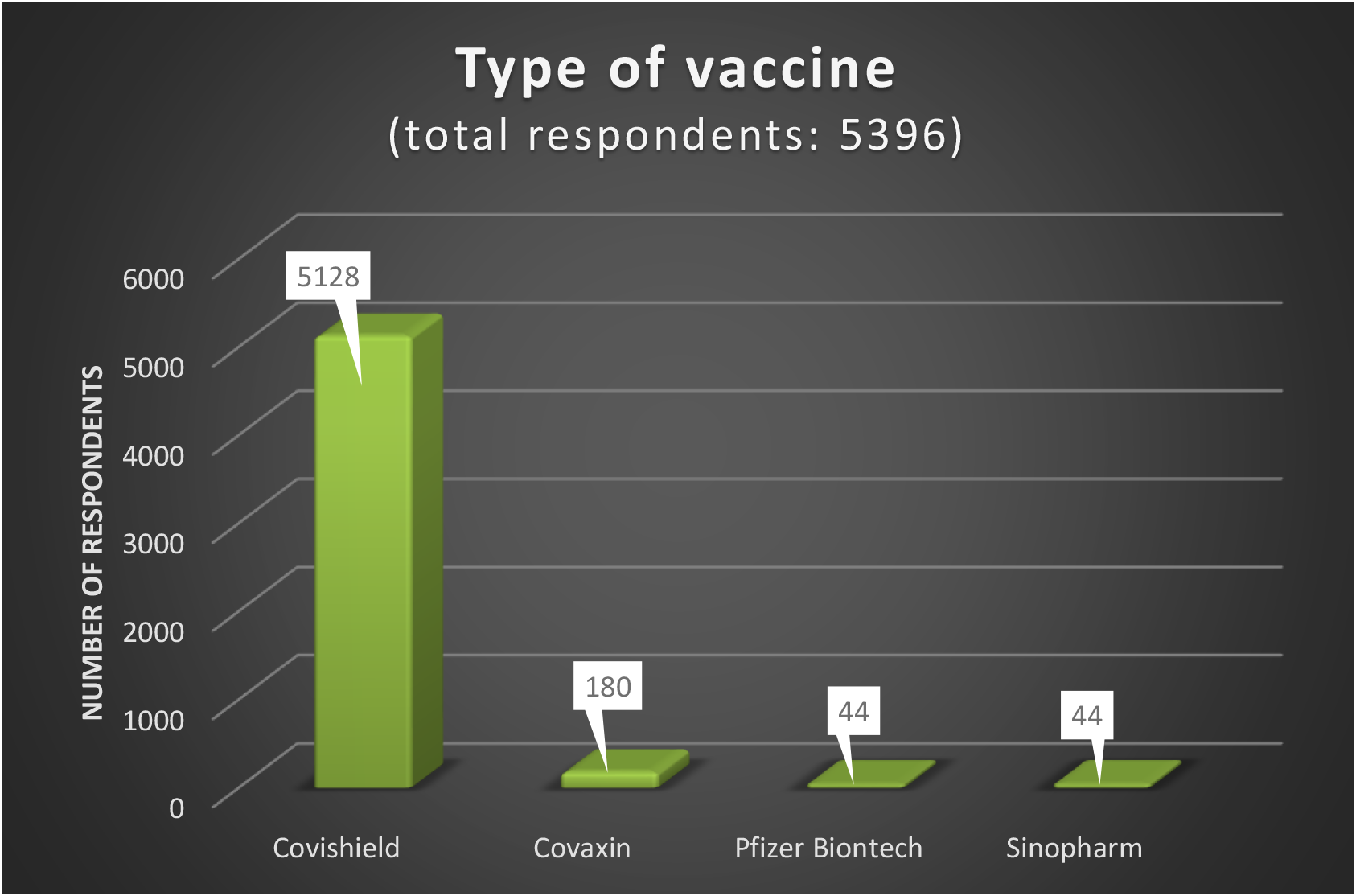
Type of vaccine.

Overall, 65.9 % of respondents reported at least one post-vaccination symptom.

Tiredness (45%), myalgia (44%), fever (34%), headache (28%), local pain at injection site (27%), joint pain (12%), nausea (8%) and diarrhoea (3%) were the most prevalent symptoms. All other symptoms were 1% or less (Figure 2).

**Figure 2:**
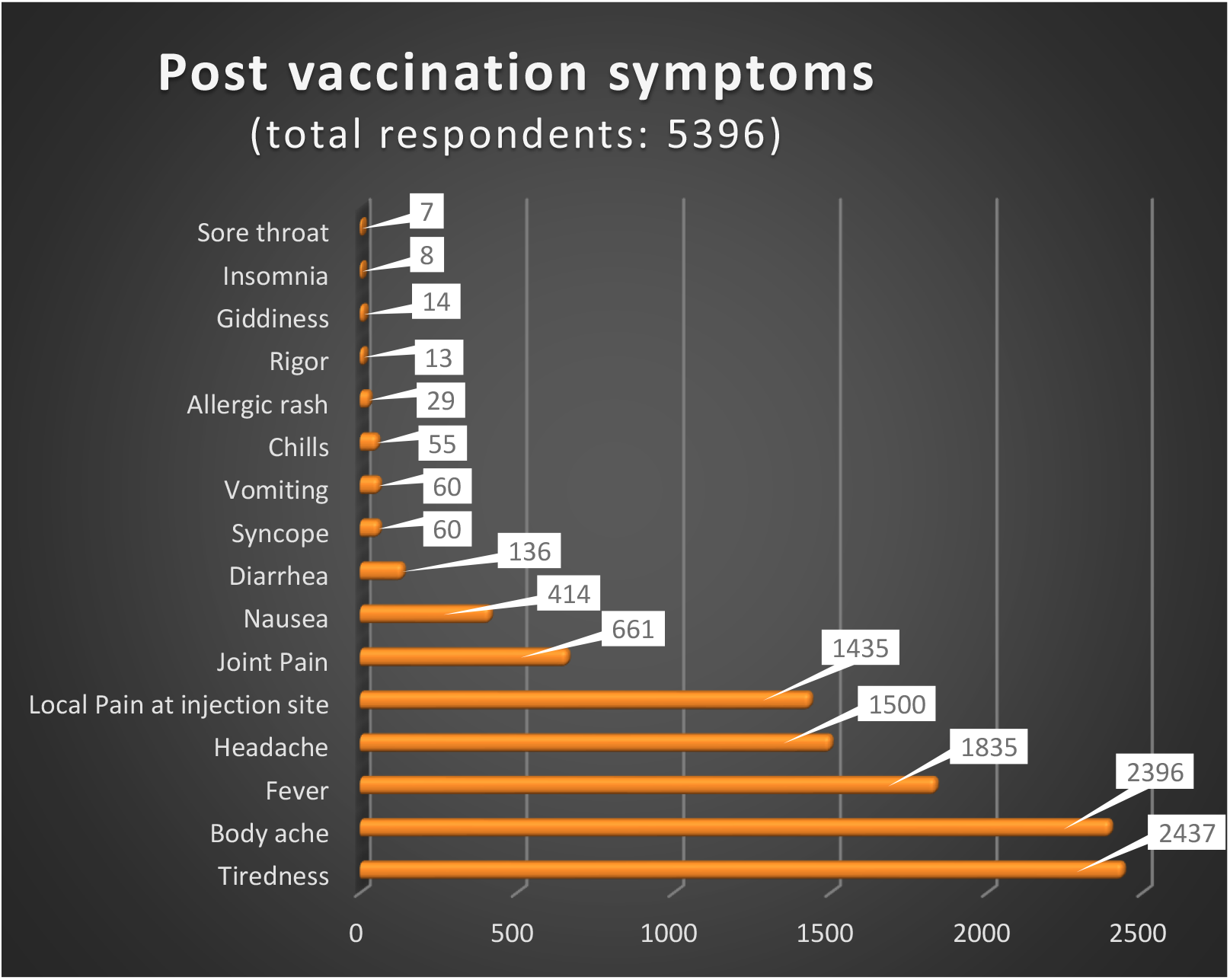
Description of post-vaccination symptoms.

The chance of having symptoms decreased with advancing age (Figure 3). The frequency of symptoms was 81% (3^rd^ decade or 20-29 years), 80% (4^th^ decade or 30-39 years), 68% (5^th^ decade), 58% (6^th^ decade), 45% (7^th^ decade), 34% (8^th^ decade) and 7% (9^th^ decade, 80-90 years). (Table 1)

**Table 1:** The incidence and severity of post-vaccination symptoms with age.

| Assessments |  | Symptoms by age group |  |  |  |  |  |  | P Value |
| --- | --- | --- | --- | --- | --- | --- | --- | --- | --- |
|  |  | 20-30 | 30-40 | 40-50 | 50-60 | 60-70 | 70-80 | 80-90 |  |
| Symptoms following vaccine | Yes | 545(81.34%) | 1122(79.57%) | 871(67.94%) | 633(58.23%) | 299(44.76%) | 85(33.73%) | 2(7.41%) | <0.001* |
|  | No | 125(18.36%) | 288(20.43%) | 411(32.06%) | 454(41.77%) | 369(55.24%) | 167(66.27%) | 25(92.59%) |  |
| Onset in hours |  | 10.05±6.23 | 10.65±6.96 | 10.96±7.19 | 12.53±7.75 | 13.39±7.83 | 13.27±8.37 | 24 | <0.001* |
| Duration in hours |  | 28.83±14.88 | 30.57±15.30 | 29.78±15.40 | 28.60±15.53 | 25.82±15.33 | 22.88±15.39 | 28.05±28.35 | <0.001* |
| Inconvenience to work | Yes | 275(41.04%) | 404(28.65%) | 227(17.71%) | 122(11.22%) | 49(7.34%) | 10(3.97%) | 0 | <0.001* |
|  | No | 395(58.96%) | 1006(71.35%) | 1055(97.50%) | 965(88.78%) | 619(72.66%) | 242(96.03%) | 27(100%) |  |
| Paracetamol Intake | Yes | 493(73.58%) | 986(69.93%) | 792(61.78%) | 569(52.35%) | 284(42.51%) | 89(35.32%) | 5(18.52%) | <0.001* |
|  | No | 177(26.42%) | 424(30.07%) | 490(38.22%) | 518(47.65%) | 384(57.49%) | 163(64.68%) | 22(81.48%) |  |

**Figure 3:**
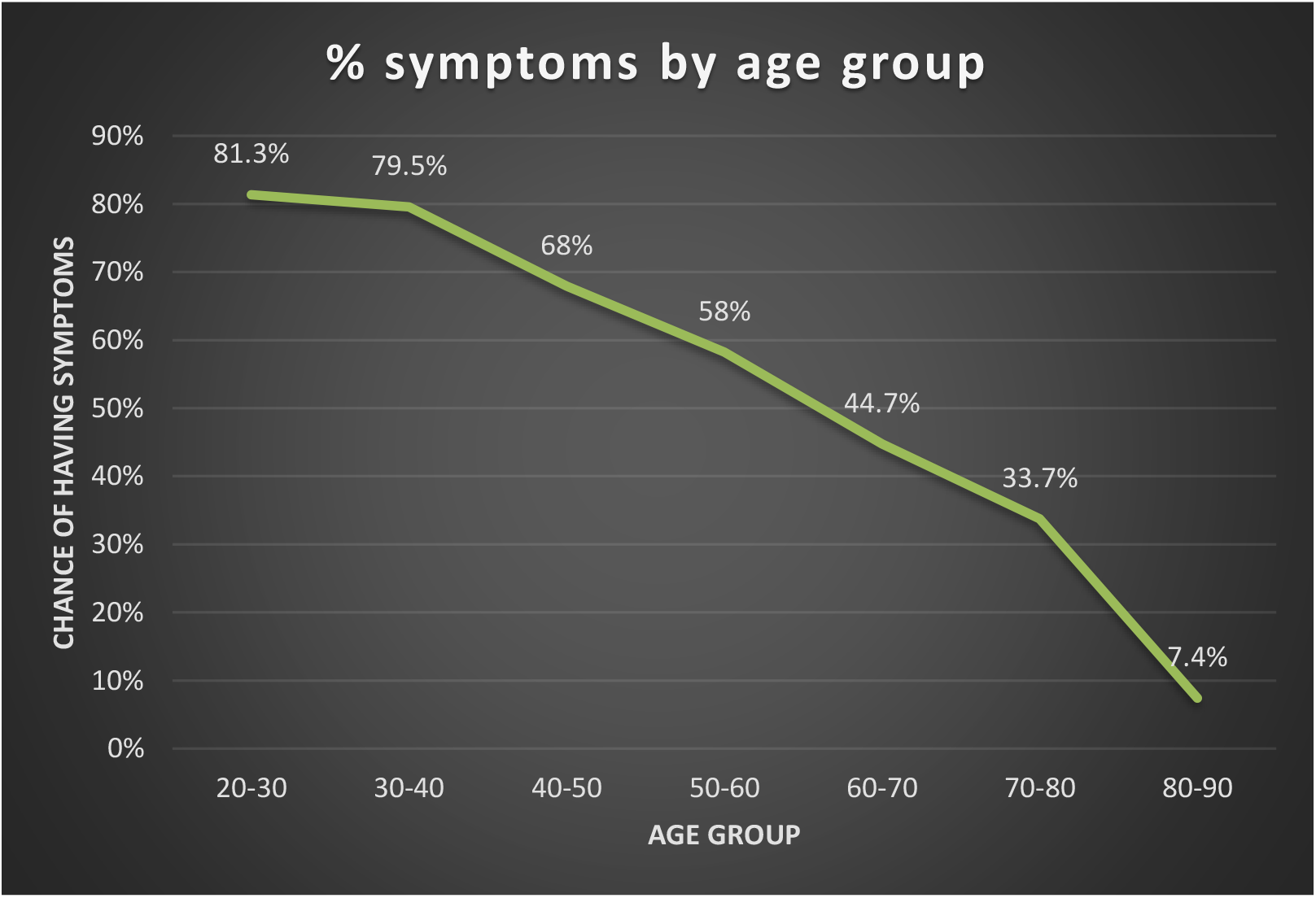
Incidence of post-vaccination symptoms by age group.

Post-vaccination symptoms were more likely to be reported by women (74.7%) compared to men (58.6%) (p < 0.001), this observation was consistent across all age groups. Women were more likely to report symptoms severe enough to prevent working for a day (27% vs. 15%) and the need to take pain relievers (70% vs. 51%). Women developed symptoms slightly earlier (10 hours after vaccination) than men (12 hours) (p < 0.001). Women had slightly longer duration of symptoms (30 hours vs. 28.5 hours, p = 0.01) (Table 2).

**Table 2:**
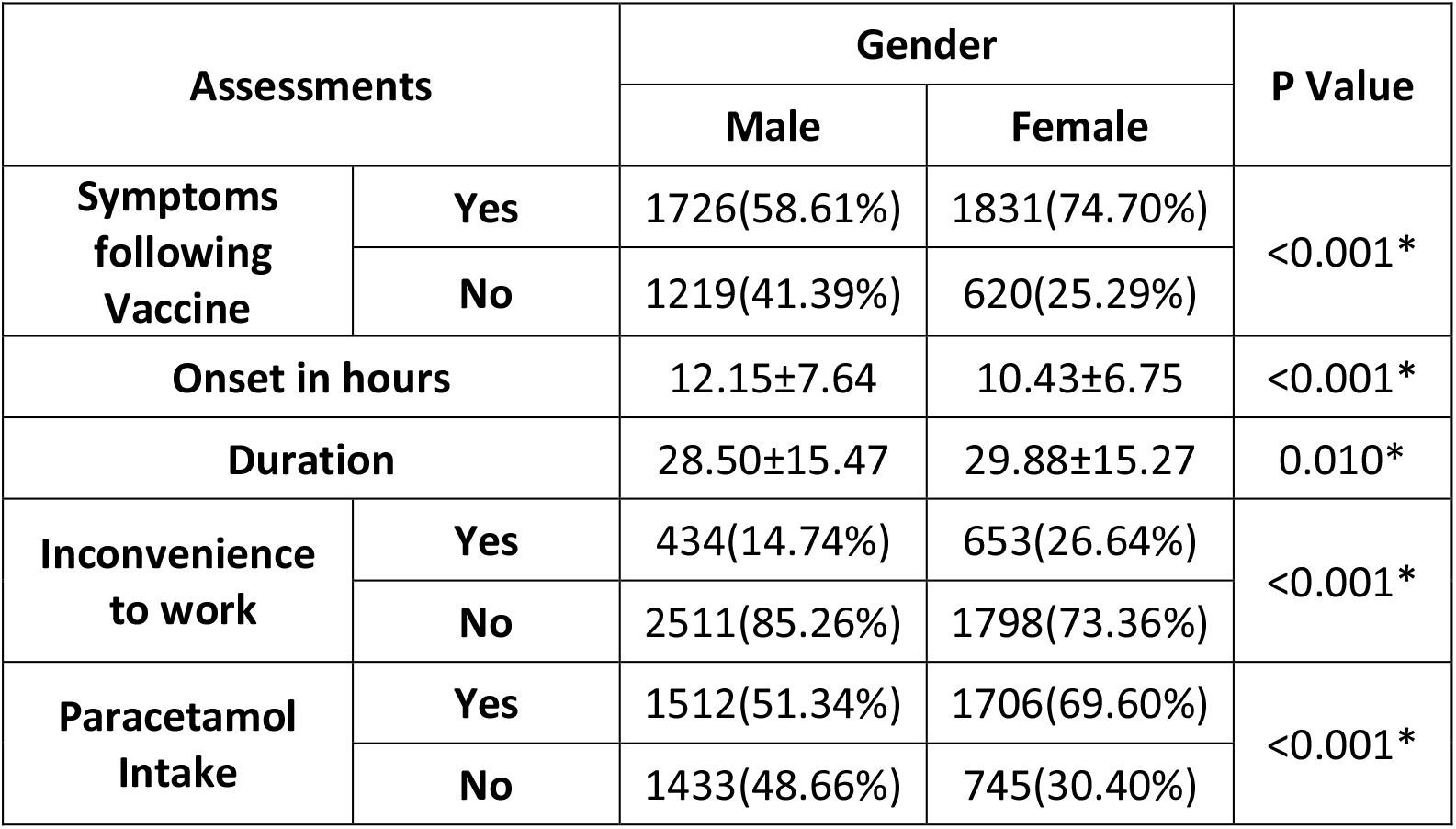
Post-vaccination symptoms among men vs. women.

Older people had later onset of symptoms, occurring at an average of 13.4 hours (70-79 years), compared to 10 hours in younger age groups (20-29 years) following vaccination. (P<0.001) The duration of symptoms decreased with advancing age, ranging from an average of 28.8 hours in younger age groups (20-29 years) to 22.9 hours in older age groups (70-79 years) (P<0.001).

Among those who reported symptoms, 79% noticed them within the first 12 hours.

472 out of 5396 (8.7%) reported past history of COVID-19. Their symptom profile was not different to those who did not have a past history. (Table 3)

**Table 3:** Post-vaccination symptoms based on past history of COVID-19.

| Symptom parameters |  | Past history of COVID-19 |  |
| --- | --- | --- | --- |
|  |  | Yes | No |
| Symptoms following Vaccine | Yes | 314(66.53%) | 3243(65.86%) |
|  | No | 158(33.47%) | 1681(34.14%) |
| Onset (hours since injection) |  | 12.31±7.58 | 11.18±7.21 |
| Duration |  | 29.67±15.84 | 29.17±15.33 |
| Inconvenience to work | Yes | 106(22.46%) | 981(19.92%) |
|  | No | 366(77.54%) | 3943(80.08%) |
| Paracetamol Intake | Yes | 279(59.11%) | 2939(59.69%) |
|  | No | 193(40.89%) | 1985(40.31%) |

The frequency of experiencing symptoms was 66.6% for Covishield (5128 responses) and 55% for Covaxin (180 responses). As only 3.3% of the respondents in this survey received Covaxin, direct comparison was not feasible. Among those who took Pfizer and Sinopharm vaccines, 70.7% and 24.4% each reported symptoms. (44 responses each) (Table 5)

The mean duration of symptoms was 30 hours (Covishield), 26 hours (Covaxin), 32 hours (Pfizer) and 20 hours (Sinopharm).

13 responses came from people who declined the vaccine. The reasons stated were:

1. Doubt regarding the duration of immunity from vaccination
2. Fear of anaphylaxis reaction
3. Fear of SARS-CoV-2 infection
4. Sceptical of the studies done so far
5. Did not believe in the quality of the vaccine
6. Fear of long-term complications

## Discussion

Healthcare professionals who took the survey provided a description of their immediate post-vaccination experience. Two-thirds of survey respondents reported mild and predictable symptoms following vaccination. The remaining one-third did not report any symptom.

Tiredness was the most common symptom (45%), followed by myalgia (44%), fever (34%) and headache (28%). Local pain at the injection site was reported by 27% of the respondents.

None of the symptoms were of serious nature or requiring hospitalization. Symptoms appeared within 11 hours (2-24 hours) after vaccination; mean duration was 30 hours. 90% reported that the post-vaccination symptom severity was as they had expected or milder. 10% reported that the symptoms were worse than they had expected. 20% felt that the symptoms were severe enough to affect work the following day.

The frequency of experiencing symptoms following each vaccine were 66% (Covishield), 53% (Covaxin), 70.7% (Pfizer) and 24.4% (Sinopharm).

There was a clear linear correlation between age and post-vaccination symptoms, suggesting that vaccine reactogenicity declined with age. In the youngest age group (20-29 years) 81.3% developed symptoms, while only 7.4% of those over 80 years reported any symptom. Vaccine reactogenicity is known to correlate with transient elevation of inflammatory cytokines, but is not considered a reliable sign of a desirable immune response (1)

Women were more likely to develop post-vaccination symptoms. The onset of symptoms was slightly earlier and the duration slightly longer in this group. This observation was consistent across all age groups.

The findings of the survey correlated with results from published trials of vaccines. In the phase 2/3 trial of Astra-Oxford ChAdOx1 nCoV-19, at least one systemic symptom was reported following vaccination with the standard dose by 86% participants in the 18–55 years group, 77% in the 56–69 years group, and 65% in the 70 years and older group (2)

While discussing post vaccination experience, it is noteworthy that placebo injections produce comparable symptoms. In the phase 3 trial of Pfizer-Biontech vaccine, the incidence of headache following vaccination was 42% in the vaccine group and 34% in those who received saline placebo (3). This has been termed the nocebo effect, which results from enhanced anticipation of negative outcomes from an intervention (4).

This study did not measure post vaccination antibody response. Hence it is not possible to infer whether the muted post-vaccination symptoms among older people was a sign of immune senescence. Although symptoms are known to correlate with neutralising antibody levels during COVID-19 (5), the presence of symptoms following vaccination does not reliably predict antibody response (6).

The frequency of using paracetamol to reduce post vaccination symptoms decreased from 71% in the 20-29 age group to 16% in the 80-90 age group. This correlated with the symptom frequency in these subgroups. Although the use of paracetamol to alleviate post-vaccination discomfort is considered acceptable (7), routine prophylactic use of pain-relievers is not recommended as there is evidence of blunted immune response as a result (7,8).

Fear of the unknown is a driver of vaccine hesitancy. By describing what to expect, the findings of this study will be reassuring to those who are fearful of the new vaccine. The fact that post-vaccination symptoms were mild, predictable and short-lived will help reduce vaccine hesitancy.

## Limitations

Any survey is more likely to be taken by those with an interest in the topic. For instance, a survey on side effects of a medication is more likely to be answered by someone who had a problem with that medication. Those who developed symptoms following vaccination could likewise show greater interest in the survey. Greater awareness and anticipation of potential adverse effects among healthcare workers as a group, could also get reflected in the reporting rate. Hence, the reported 65% incidence of post-vaccination symptoms could be an overestimation.

Survey questions were in English, which might have posed some difficulty among those with limited proficiency in that language.

As the survey was done soon after initiation of vaccination, there was no scope for reporting delayed symptoms. The survey was done on trust; it was not possible to verify the I.D or information provided by each respondent. The relatively small number of respondents who took the Covaxin, Pfizer and Sinopharm vaccines makes it difficult to do a head to head comparison.

## Conclusions

Two-thirds of healthcare professionals who completed the survey reported mild and short-lived post-vaccination symptoms. Tiredness, myalgia and fever were most commonly reported. These symptoms were consistent with an immune response commonly associated with vaccines, and correlated with the findings from previously published phase 2/3 trials. In 90% cases, the symptoms were either milder than expected or meeting the expectation of the vaccine recipient. No serious events were reported. Symptoms were more common among younger individuals. There was no difference in symptoms among those who had a past history of COVID-19.

## Data Availability

Data will be furnished upon request, including the raw data excel file of the 5396 respondents.

